# Three-dimensional pulp chamber volume quantification in first molars using CBCT: Implications for machine learning-assisted age estimation

**DOI:** 10.1101/2025.08.05.25333098

**Authors:** Yanjie Ding, Tao Zhong, YuXin He, Wei Wang, Shilin Zhang, Xiao Zhang, WenLi Shi, Bo Jin

## Abstract

Accurate adult age estimation represents a critical component of forensic individual identification. However, traditional methods relying on skeletal developmental characteristics are susceptible to preservation status and developmental variation. Teeth, owing to their exceptional taphonomic resistance and minimal postmortem alteration, emerge as premier biological samples. Utilizing the high-resolution capabilities of Cone Beam Computed Tomography (CBCT), this study retrospectively analyzed 1,857 right first molars obtained from Han Chinese adults in Sichuan Province (883 males, 974 females; aged 18–65 years). Pulp chamber volume (PCV) was measured using semi-automatic segmentation in Mimics software (v21.0). Statistically significant differences in PCV were observed based on sex and tooth position (maxillary vs. mandibular). Significant negative correlations existed between PCV and age (r = -0.86 to -0.81). The strongest correlation (r = -0.88) was identified in female maxillary first molars. Eleven curvilinear regression models and six machine learning models (Linear Regression, Lasso Regression, Neural Network, Random Forest, Gradient Boosting, and XGBoost) were developed. Among the curvilinear regression models, the cubic model demonstrated the best performance, with the female maxillary-specific model achieving a mean absolute error (MAE) of 4.95 years. Machine learning models demonstrated superior accuracy. Specifically, the sex- and tooth position-specific XGBoost model for female maxillary first molars achieved an MAE of 3.14 years (R² = 0.87). This represents a significant 36.5% reduction in error compared to the optimal cubic regression model. These findings demonstrate that PCV measurements in first molars, combined with machine learning algorithms (specifically XGBoost), effectively overcome the limitations of traditional methods, providing a highly precise and reproducible approach for forensic age estimation.

## 1. Introduction

Age estimation represents a fundamental task in forensic individual identification and plays a critical role in judicial contexts, including victim identification in mass disaster scenarios, criminal investigations, and age verification in immigration cases [1, 2]. Traditional methods primarily rely on skeletal developmental characteristics, such as cranial suture closure, pubic symphysis morphology, and the degree of epiphyseal fusion in long bones. However, these methods are susceptible to preservation status and developmental variation, particularly in skeletonized remains or mass disaster scenarios, resulting in significant estimation errors averaging 5 years or more [3–5]. Conversely, teeth represent superior biological specimens for age estimation owing to their excellent taphonomic resistance, conferring resilience against decomposition, high temperatures, and mechanical trauma. Additionally, dental development exhibits reduced susceptibility to interference from nutritional, metabolic, and pathological factors [6].

Age-related alterations in the pulp chamber arise primarily from the continuous deposition of secondary dentin [7]. Following Gustafson’s initial proposal of an association between this process and age [8], Philippas et al pioneered the application of radiographic imaging to assess pulp chamber morphology, enabling the utilization of one-dimensional parameters [9], including root length and pulp chamber width [10, 11]. Subsequent investigations explored two-dimensional parameters [12], such as the pulp to tooth area ratio. However, these parameters demonstrated limited correlation with age and proved insufficient to meet the precision demands of forensic applications. This limitation arises from the inherently complex three-dimensional geometry of the pulp chamber, the morphology of which is affected by tooth rotation and the uneven deposition of secondary dentin. Conventional two-dimensional radiographic techniques, compromised by projection distortion and insufficient density resolution, are unable to accurately quantify these three-dimensional morphological changes.

With the advancement of three-dimensional imaging technologies, particularly the introduction of cone beam computed tomography (CBCT) into forensic odontology, a significant advance has been provided in individual identification [13]. This technology employs a cone-shaped X-ray beam coupled with high-sensitivity detectors (achieving voxel sizes ≤200 μm) to reconstruct high-resolution three-dimensional images with slice thicknesses below 0.1 mm, enabling precise visualization of morphological details within the dentino-pulpal complex. For example, Yangjing S. et al. established a logarithmic regression model based on pulp chamber morphology in first molars, achieving a mean absolute error (MAE) of 6.72 years [14]. Compared with conventional two-dimensional imaging, CBCT offers several key advantages: (1) Elimination of anatomical structure superimposition, enabling direct volumetric quantification of the pulp chamber; (2) Reduction of contour recognition errors induced by tooth inclination or pulp cavity calcification; (3) Delivery of a substantially reduced radiation dose, aligning with the ethical and safety standards of forensic practice. However, current CBCT-based age estimation studies primarily utilize metrics such as pulp cavity volume or tooth volume [15]. Furthermore, research indicates that occlusal enamel wear and root-related factors may compromise estimation accuracy [16, 17]. The first permanent molar, owing to its early eruption, high functional load, and distinct CBCT-visualized anatomical landmarks, presents considerable potential for forensic application [18]. Therefore, this study employed CBCT technology to develop an age estimation model utilizing pulp chamber volume (PCV) in first molars as the primary metric. This parameter effectively minimizes interference from occlusal enamel wear and excludes the influence of root-related factors, thereby enhancing both methodological feasibility and result reliability.

However, traditional CBCT-based methodologies for measurement and age estimation possess inherent limitations that hinder their capacity to satisfy the rigorous precision demands of forensic practice. Consequently, machine learning (ML) methods have gained significant traction in the medical field. A key advantage of ML lies in its ability to model complex non-linear relationships and identify latent patterns in data [19, 20]. Studies utilizing dental morphological data from conventional radiographs [21, 22] for dental age estimation (DAE) have demonstrated that incorporating ML techniques can substantially improve accuracy compared to traditional formula-based approaches. This data-driven methodology, characterized by automated feature extraction, presents a promising framework for establishing a standardized, high-precision system for forensic dental age estimation.

Therefore, this study investigated a Han Chinese adult population from Sichuan Province, China. Utilizing three-dimensional CBCT image data, PCV of first molars was measured to develop and compare curvilinear regression models and machine learning models for age estimation. This comparative analysis aimed to offer a precise and reproducible method for forensic age estimation.

## 2. Materials and methods

### 2.1. Study Population

CBCT image data were retrospectively collected from the dental database of the Affiliated Hospital of North Sichuan Medical College between December 2020 and June 2023. All images were stored in Digital Imaging and Communications in Medicine (DICOM) format.

Inclusion criteria were as follows:

1. Han Chinese individuals aged 18–65 years;
2. Absence of dental caries, excessive occlusal wear, restorations, or pulp calcification in the target teeth;
3. CBCT images depicting a fully intact and clearly discernible right first molar.

Exclusion criteria included:

1. Age outside the 18–65-year range;
2. Pathology (e.g., disease, trauma, developmental malformations) or prior surgical intervention involving the right first molar;
3. History of medication use known to affect dental metabolism, development, or growth;
4. Presence of systemic conditions affecting dental growth, development, or metabolism;
5. Incomplete personal information.

The study sample comprised retrospectively collected CBCT images obtained during routine clinical dental diagnosis and treatment. The collection and analysis of these data complied with the principles of the Declaration of Helsinki. The study protocol was reviewed and granted a waiver of informed consent by the Institutional Review Board (IRB) of North Sichuan Medical College (Approval No.: 2023006).

### 2.2. Data Acquisition and Collection

A total of 1,857 CBCT scans (883 from males, 974 from females) were collected from Han Chinese individuals aged 18–65 years residing in Sichuan Province, China. All CBCT scans were acquired using an OP 3D Vision CBCT system (KaVo Dental GmbH, Biberach, Germany). The scanning parameters were as follows: tube voltage, 120 kV; tube current, 5 mA; exposure time, ∼4 s; slice thickness, 0.2 mm; window level, 600 HU; window width, 3200 HU. For each case, sex, date of birth (DOB), and scan date were recorded. Chronological age at the time of scanning was calculated by subtracting the DOB from the scan date. The age distribution and corresponding sample sizes for the study cohort are presented in Table 1.

**Table 1.** Age and sex distribution of the study subjects.

| Age Group | Males | Females | Total |
| --- | --- | --- | --- |
| [18, 27) | 169 | 201 | 370 |
| [28, 37) | 267 | 233 | 500 |
| [38, 47) | 230 | 254 | 484 |
| [48, 57) | 175 | 222 | 397 |
| [58, 65] | 42 | 64 | 106 |
| Total | 883 | 974 | 1857 |

### 2.3. PCV Measurement

Blinded to the chronological age and sex of the individuals, two observers, trained uniformly in the measurement protocol, measured the pulp chamber volume (PCV) of the right maxillary (tooth 16) and mandibular (tooth 46) first molars for all cases using CBCT image data.

To assess inter- and intra-observer reliability, a preliminary study was conducted. Both observers independently measured PCV in a randomly selected subset of 200 CBCT scans. The same subset was re-measured by both observers after a four-week interval to assess intra-observer reliability.

PCV measurement was performed using Mimics software (v21.0; Materialise NV, Leuven, Belgium) according to the following workflow:

(1) DICOM files were imported into Mimics software.

(2) The target teeth (teeth 16 and 46) were identified and isolated in the three-dimensional (3D) reconstruction view.

(3) Segmentation was initiated using the ‘SEGMENT’ tab. Key functionalities employed were:

Thresholding: This function applies a defined Hounsfield Unit (HU) range to create a binary mask, retaining voxels within the specified upper and lower limits.

Region Growing: Starting from a user-defined seed point within the pulp chamber region, this function iteratively adds connected voxels meeting the threshold criteria to the mask, extracting contiguous structures.

(4) Based on the lower Hounsfield Unit (HU) values characteristic of pulp tissue compared to surrounding dentin, an appropriate HU threshold range was applied. This enabled semi-automatic segmentation of the pulp chamber primarily using the thresholding and region growing functions described in steps (3).

(5) Due to frequent imprecision in the automatically generated masks, manual refinement was performed on the initial mask using editing tools. Specific functions used for refinement included ‘Edit Masks’ and ‘Multiple Slice Edit’, allowing for manual addition or removal of extraneous voxels.

(6) Following final modifications, the refined mask representing the pulp chamber was selected. The ‘Calculate Part’ function was then executed, generating a three-dimensional (3D) surface model of the pulp chamber (Fig 1).

**Fig. 1.**
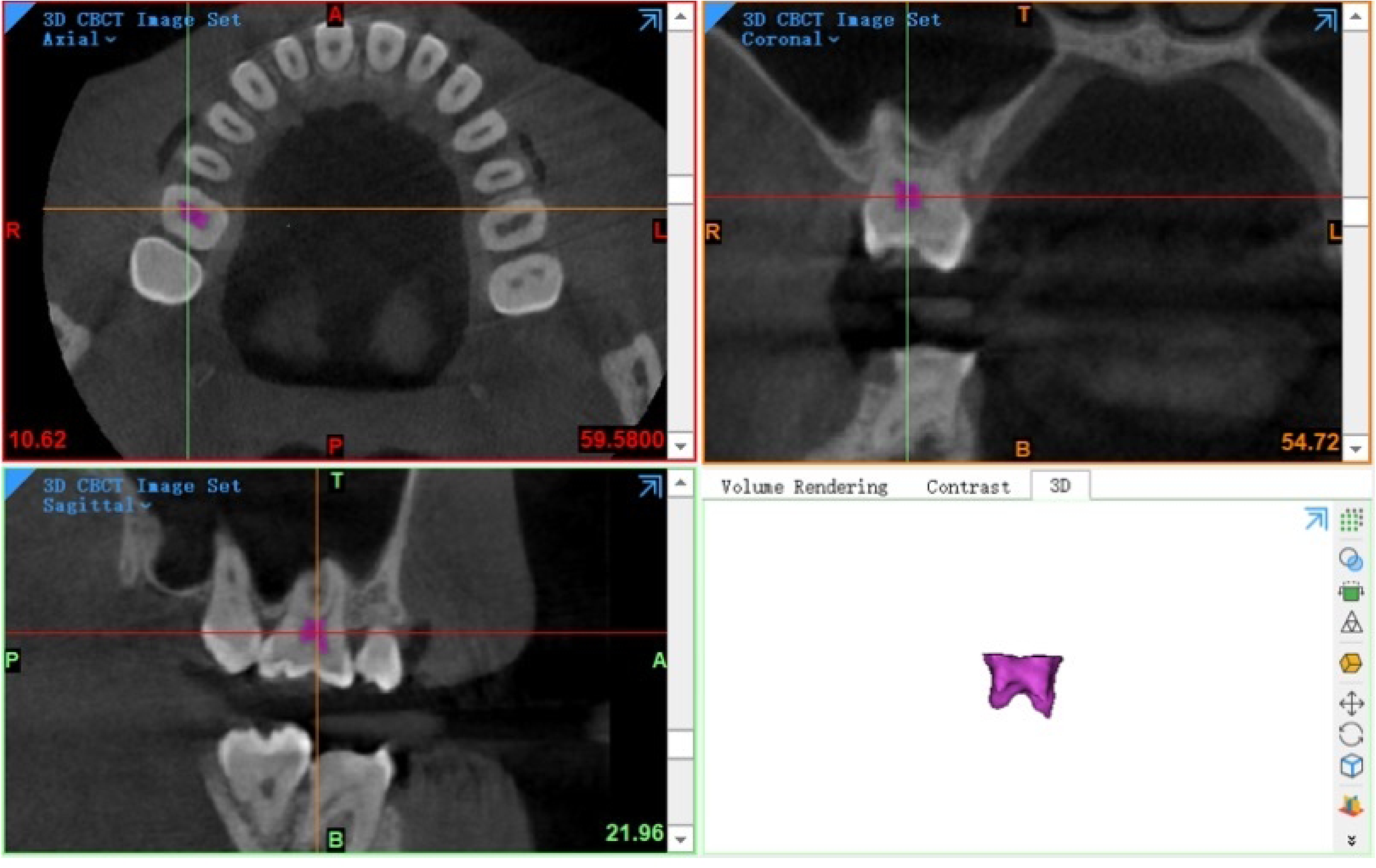
Visualization of 3D pulp chamber segmentation.

(7) For both tooth 16 and tooth 46, the final pulp chamber mask was converted into a 3D model.

(8) For each generated 3D pulp chamber model, the ‘Properties’ option was selected from the menu, and the volume value (mm³) displayed in the dialog box was recorded.

### 2.4. Age Estimation Model Development

Chronological age served as the dependent variable (*y*), and pulp chamber volume (PCV) as the independent variable (*x*). Eleven traditional regression models were developed using the training set (n=1,485): linear, logarithmic, inverse, quadratic, cubic, compound, power, S-curve, growth, exponential, and logistic regression.

Six machine learning (ML) models for age estimation were developed using the Python programming language (v3.11.6) with key libraries including NumPy, pandas, scikit-learn, and Matplotlib for data handling and model implementation: Linear Regression, Lasso Regression, Neural Network (NN), Random Forest (RF), Gradient Boosting (GB), and eXtreme Gradient Boosting (XGBoost).The source dataset comprised PCV measurements from all samples.

This source dataset was randomly partitioned into a training set (80%) and an independent test set (20%). Model hyperparameters were optimized using k-fold cross-validation (k=5) within the training set. This procedure mitigates the risk of overfitting to the specific training data partition used in cross-validation.The final performance of the optimized ML models was evaluated on the independent test set. Model performance was quantified using the mean absolute error (MAE) and coefficient of determination (R²).

### 2.5 Statistical analysis

Statistical analyses were conducted using Microsoft Excel 2021, SPSS (v25.0; IBM Corp., Armonk, NY, USA), and Python (v3.11.6). The significance level (α) was set at 0.05 for all statistical tests. Intra- and inter-observer reliability were quantified using the intraclass correlation coefficient (ICC). Model performance was evaluated using the following metrics: Coefficient of determination (R²); Root mean square error (RMSE); Mean absolute error (MAE). R² represents the proportion of variance in chronological age explained by the model, ranging from 0 to 1. Higher R² values indicate better model fit. MAE and RMSE quantify the discrepancy between predicted and actual chronological age. Lower values indicate higher prediction accuracy for both metrics.

## 3. Results

### 3.1. Reliability Assessment

Intra- and inter-observer reliability of PCV measurements were assessed using ICC. Excellent reliability was demonstrated: inter-observer ICC = 0.84, intra-observer ICC = 0.89. Both ICC values were significantly greater than zero (P < 0.05).

### 3.2. Differences in PCV by Sex and Jaw Position

Statistically significant differences (P < 0.05) were found in pulp chamber volume (PCV) between sexes and between jaw positions (maxillary vs. mandibular). Mean PCV was significantly larger in males than in females (P < 0.05; Table 2). Additionally, mean PCV was significantly larger in maxillary first molars (tooth 16) than in mandibular first molars (tooth 46) (P < 0.05; Table 2). The mean PCV across both tooth positions (16 and 46), designated as ‘Combined’ in Table 2, is also presented.

**Table 2.** PCV (mm³) in right first molars by sex and jaw position (mean ± SD).

| Tooth Position | Overall Sample(n=1857) | Males(n=883) | Females(n=974) |
| --- | --- | --- | --- |
| 16 (Maxillary) | 21.05±6.79 | 22.19±6.86 | 20.02±6.57<sup>a</sup> |
| 46 (Mandibular) | 18.25±6.13<sup>b</sup> | 19.17±6.08<sup>b</sup> | 17.43±6.07<sup>a,b</sup> |
| 16+46 (Combined) | 19.65±6.62 | 20.68±6.66 | 18.17±6.45<sup>a</sup> |

Notes: FDI tooth notation: 16 (maxillary right first molar), 46 (mandibular right first molar). The Combined represents the mean PCV across both tooth positions (16 and 46). a: Significantly different from Male group ( $P < 0.05$ ). b: Significantly different from maxillary first molar (tooth 16) ( $P < 0.05$ ).

### 3.3. Correlation Between PCV and Age

Pearson correlation analysis revealed strong negative correlations between pulp chamber volume (PCV) and chronological age (r = -0.81 to -0.88; all P < 0.05; see Table 3). The strongest negative correlation was found for the maxillary right first molar (tooth 16, FDI notation) in females (r = -0.88; P < 0.05). Across all groups, correlations for the mandibular right first molar (tooth 46, FDI notation) were consistently strong but slightly weaker than those for tooth 16. The ‘Combined’ correlation coefficient (Table 3) represents the analysis using PCV data pooled from both tooth positions (16 and 46) for each individual.

**Table 3.** Pearson correlation coefficients (r) between PCV and chronological age.

| Tooth Position | Overall Sample(n=1857) | Males(n=883) | Females(n=974) |
| --- | --- | --- | --- |
| 16 (Maxillary) | -0.86* | -0.85* | -0.88* |
| 46 (Mandibular) | -0.82* | -0.82* | -0.83* |
| 16+46 (Combined) | -0.81* | -0.81* | -0.84* |

Notes: FDI tooth notation: 16 (maxillary right first molar), 46 (mandibular right first molar). The Combined represents the Pearson correlation coefficient calculated using PCV data pooled from both tooth positions (16 and 46) for each individual. \* $p < 0.05$ .

### 3.4. Regression Model Development and Performance

Eleven curvilinear regression models were developed using pulp chamber volume (PCV) from training set subgroups: overall (n=1,485), male (n=706), and female (n=706). For each subgroup, models were built using PCV from the maxillary right first molar (tooth 16, FDI notation), the mandibular right first molar (tooth 46, FDI notation), and the mean PCV calculated from both teeth 16 and 46 for each individual (designated as Combined PCV).

The cubic regression model yielded the highest coefficient of determination (R²) among all models for each subgroup (Table 4). Among these cubic models:

**Table 4.** Goodness-of-fit (R²) of regression models for age estimation using PCV.

| Models | Overall Sample (n=1485) |  |  | Male (n=706) |  |  | Female (n=706) |  |  |
| --- | --- | --- | --- | --- | --- | --- | --- | --- | --- |
|  | 16<br>(Max) | 46<br>(Mand) | 16+46<br>(Comb) | 16<br>(Max) | 46<br>(Mand) | 16+46<br>(Comb) | 16<br>(Max) | 46<br>(Mand) | 16+46<br>(Comb) |
| Linear | 0.74 | 0.68 | 0.68 | 0.73 | 0.67 | 0.66 | 0.77 | 0.701 | 0.69 |
| Logarithmic | 0.76 | 0.71 | 0.70 | 0.76 | 0.70 | 0.69 | 0.79 | 0.73 | 0.73 |
| Inverse | 0.71 | 0.68 | 0.66 | 0.71 | 0.68 | 0.66 | 0.74 | 0.69 | 0.68 |
| Quadratic | 0.77 | 0.71 | 0.71 | 0.77 | 0.71 | 0.70 | 0.79 | 0.73 | 0.73 |
| **Cubic** | **0.78** | **0.72** | **0.72** | **0.79** | **0.73** | **0.72** | **0.80** | **0.74** | **0.75** |
| Compound | 0.74 | 0.69 | 0.68 | 0.73 | 0.68 | 0.67 | 0.77 | 0.71 | 0.71 |
| Power | 0.73 | 0.69 | 0.67 | 0.72 | 0.68 | 0.67 | 0.75 | 0.71 | 0.70 |
| S-curve | 0.64 | 0.62 | 0.60 | 0.64 | 0.62 | 0.60 | 0.67 | 0.64 | 0.62 |
| Growth | 0.74 | 0.69 | 0.68 | 0.73 | 0.68 | 0.67 | 0.77 | 0.71 | 0.71 |
| Exponential | 0.74 | 0.69 | 0.68 | 0.73 | 0.68 | 0.67 | 0.77 | 0.71 | 0.71 |
| Logistic | 0.74 | 0.69 | 0.68 | 0.73 | 0.68 | 0.67 | 0.77 | 0.71 | 0.71 |

Notes: FDI tooth notation: 16 (maxillary right first molar), 46 (mandibular right first molar), 16+46 (combined PCV). Combined PCV represents the mean value of PCV from tooth 16 and tooth 46 for each individual. All values are R<sup>2</sup>. Cubic model R<sup>2</sup> values are highlighted in bold.

The female-specific model utilizing tooth 16 PCV demonstrated the lowest mean absolute error (MAE) of 4.95 years. This model is defined by:

Predicted age (y) = 79.18 - 1.981(PCV) - 0.021(PCV)² + 0.001(PCV)³

where PCV is pulp chamber volume in mm³.

Conversely, the overall model using Combined PCV yielded the highest MAE of 7.43 years:

Predicted age (y) = 82.79 - 3.023(PCV) + 0.036(PCV)² - 0.0002(PCV)³.

### 3.5. Machine Learning Model Development and

#### Performance

Building upon the observed significant variations in first molar PCV between sexes and jaw positions (Section 3.2), six machine learning models were developed: Linear Regression, Lasso Regression, Neural Network (NN), Random Forest (RF), Gradient Boosting (GB), and eXtreme Gradient Boosting (XGBoost). These models incorporated specific tooth position (maxillary vs. mandibular) and/or sex information.

Evaluation of the six ML algorithms revealed the following key findings:

Position-specific models (distinguishing maxillary vs. mandibular molars) demonstrated significantly better performance than position-agnostic models (using combined PCV data) across all sample groups ( Table 6).

In the overall sample (sex-independent): Best position-agnostic model (Gradient Boosting): R² = 0.74, MAE = 4.70 years (Fig 2) ; Position-specific XGBoost model: R² = 0.84, MAE = 3.46 years (Fig 3).

**Fig. 2.**
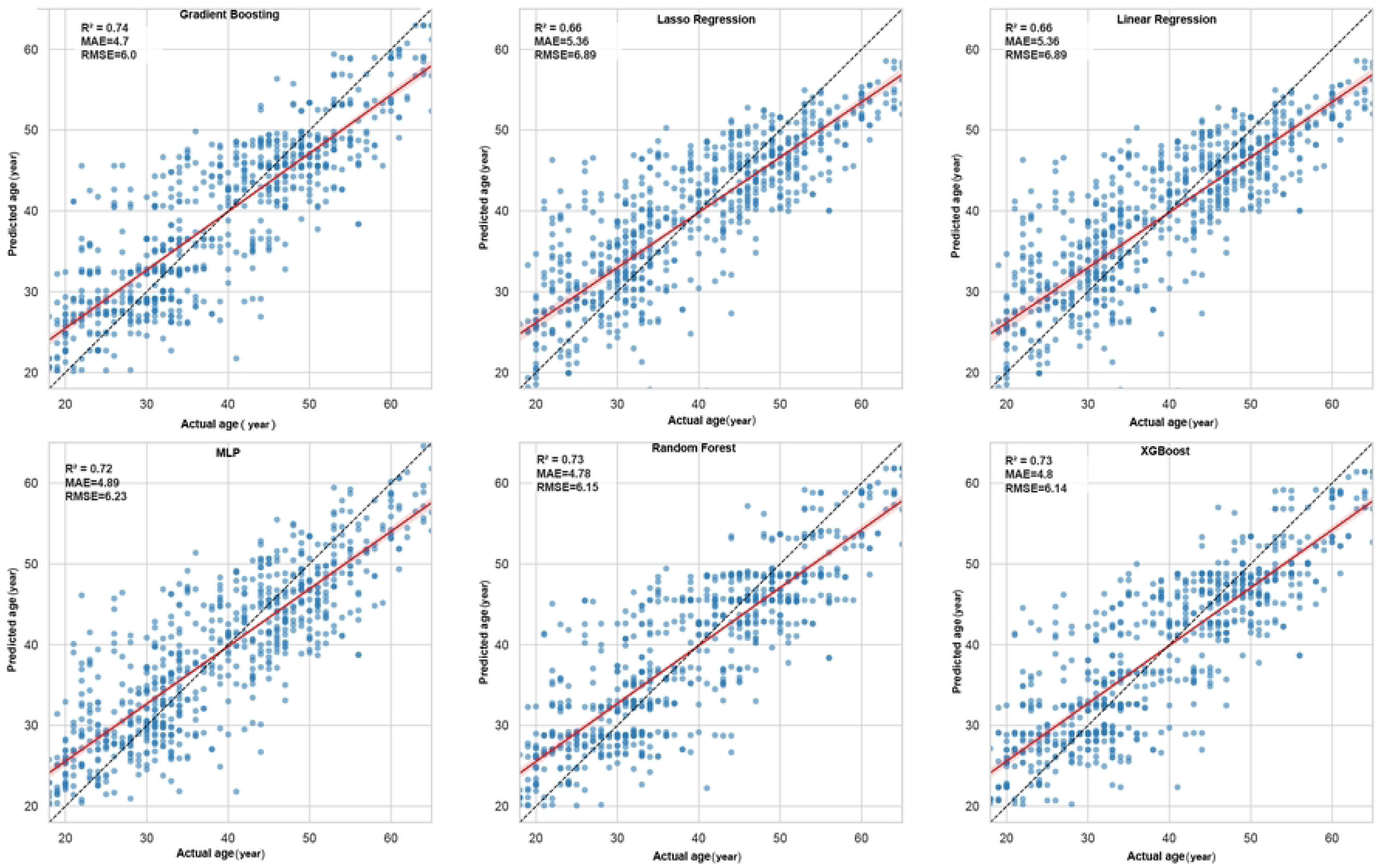
Actual vs. predicted age scatter plots for the six machine learning models in the overall sample (position-agnostic and sex-independent models).

**Fig. 3.**
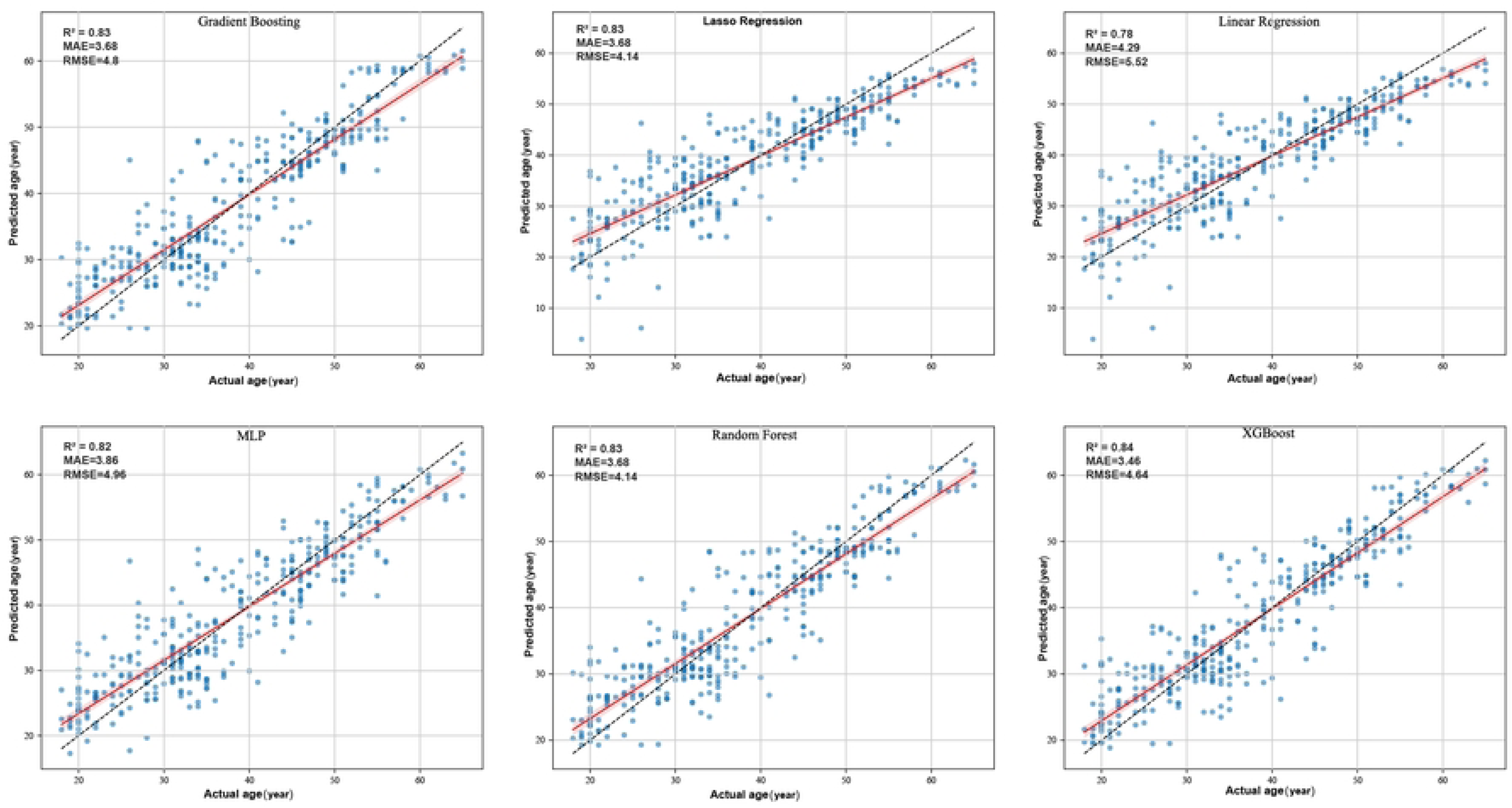
Actual vs. predicted age scatter plots for the six machine learning models in the overall sample (position-specific but sex-independent models).

In the male sample (sex-specific): Best position-agnostic model (Gradient Boosting): R² = 0.70, MAE = 4.81 years (Fig 4) ; Position-specific XGBoost model: R² = 0.83, MAE = 3.27 years (Fig 5).

**Fig. 4.**
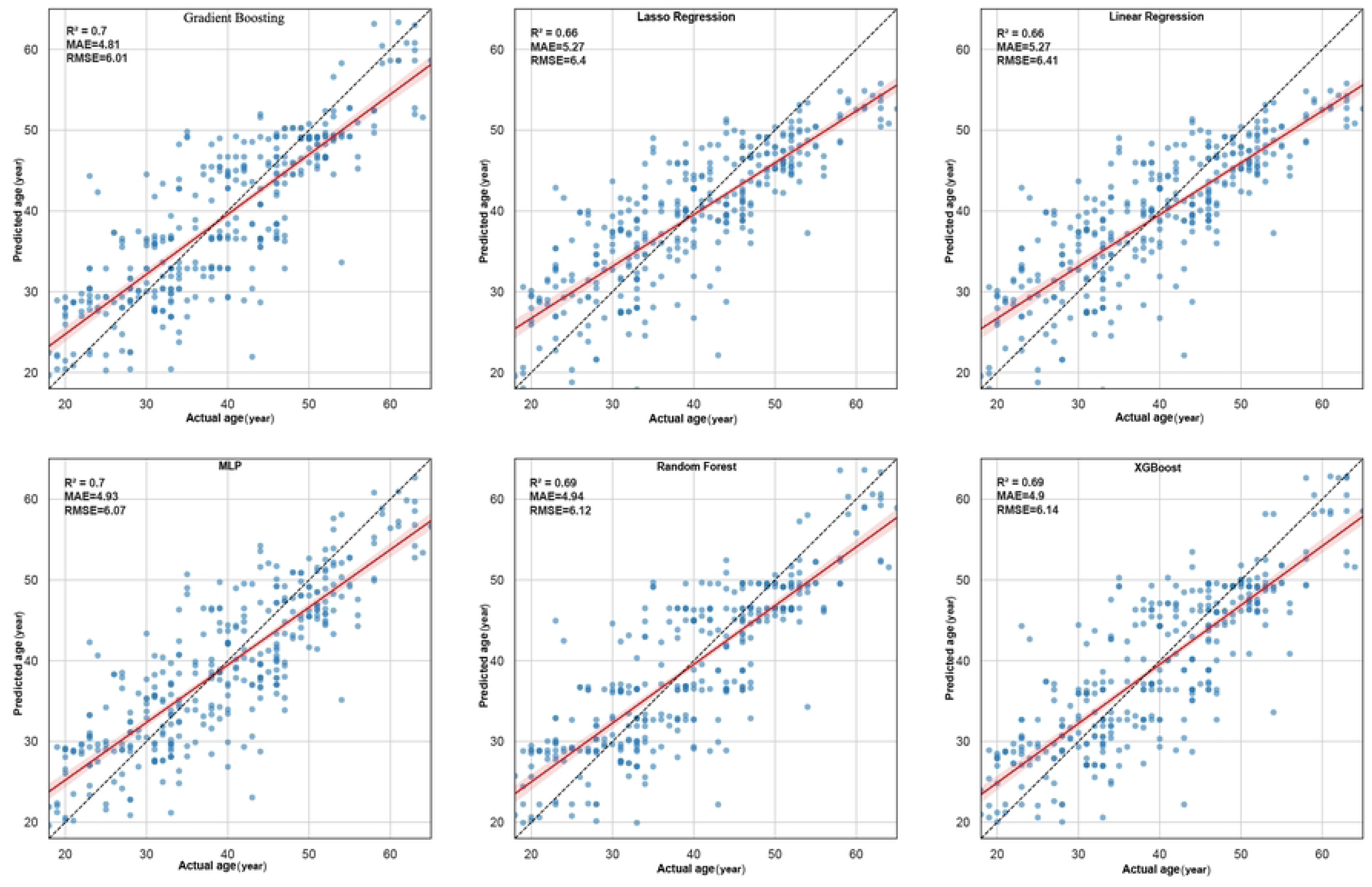
Actual vs. predicted age scatter plots for the six machine learning models in the male sample (position-agnostic but sex-specific models).

**Fig. 5.**
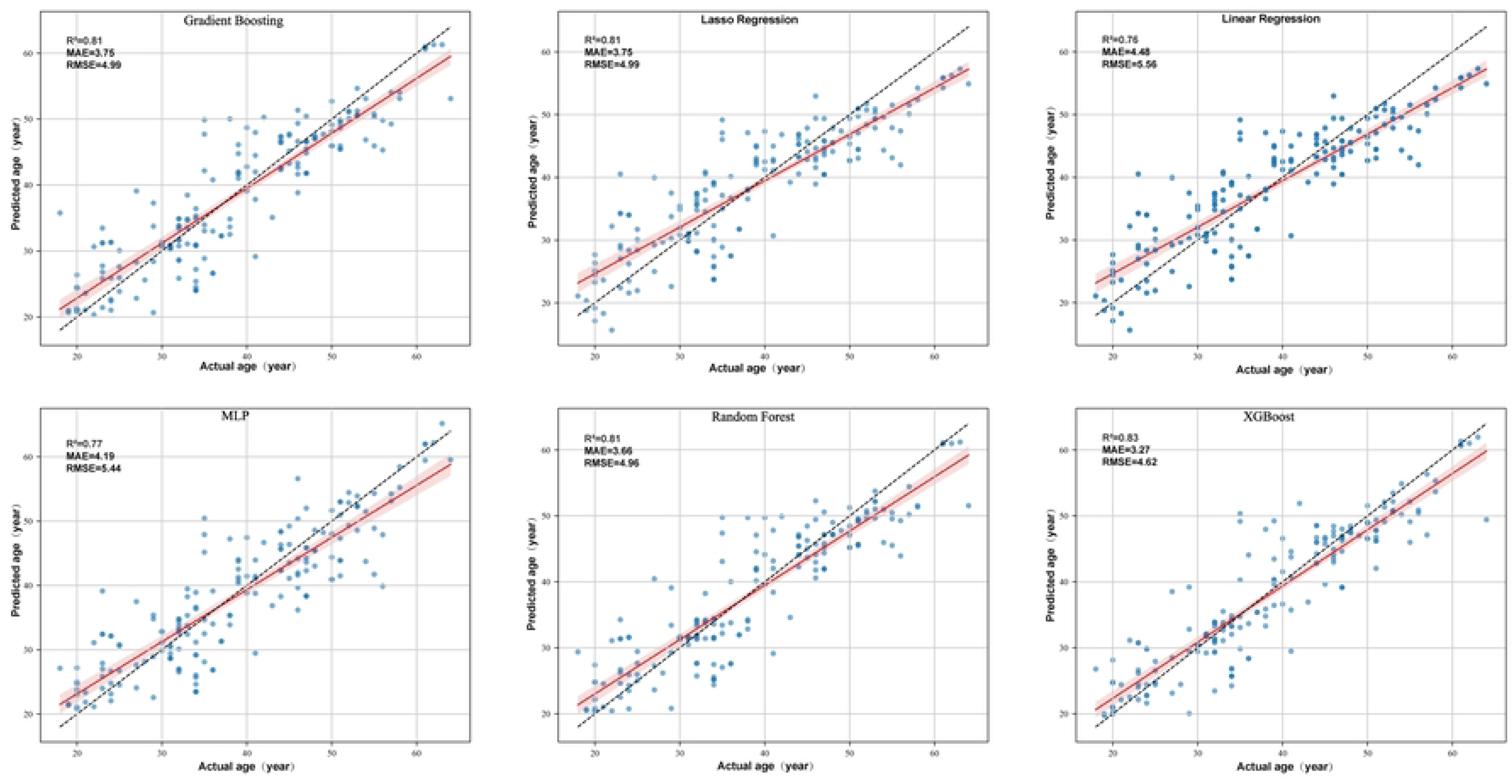
Actual vs. predicted age scatter plots for the six machine learning models in the male sample (position-specific and sex-specific models).

In the female sample (sex-specific): Best position-agnostic model (Gradient Boosting): R² = 0.74, MAE = 4.60 years (Fig 6) ; Position-specific XGBoost model: R² = 0.87, MAE = 3.14 years (Fig 7).

**Fig. 6.**
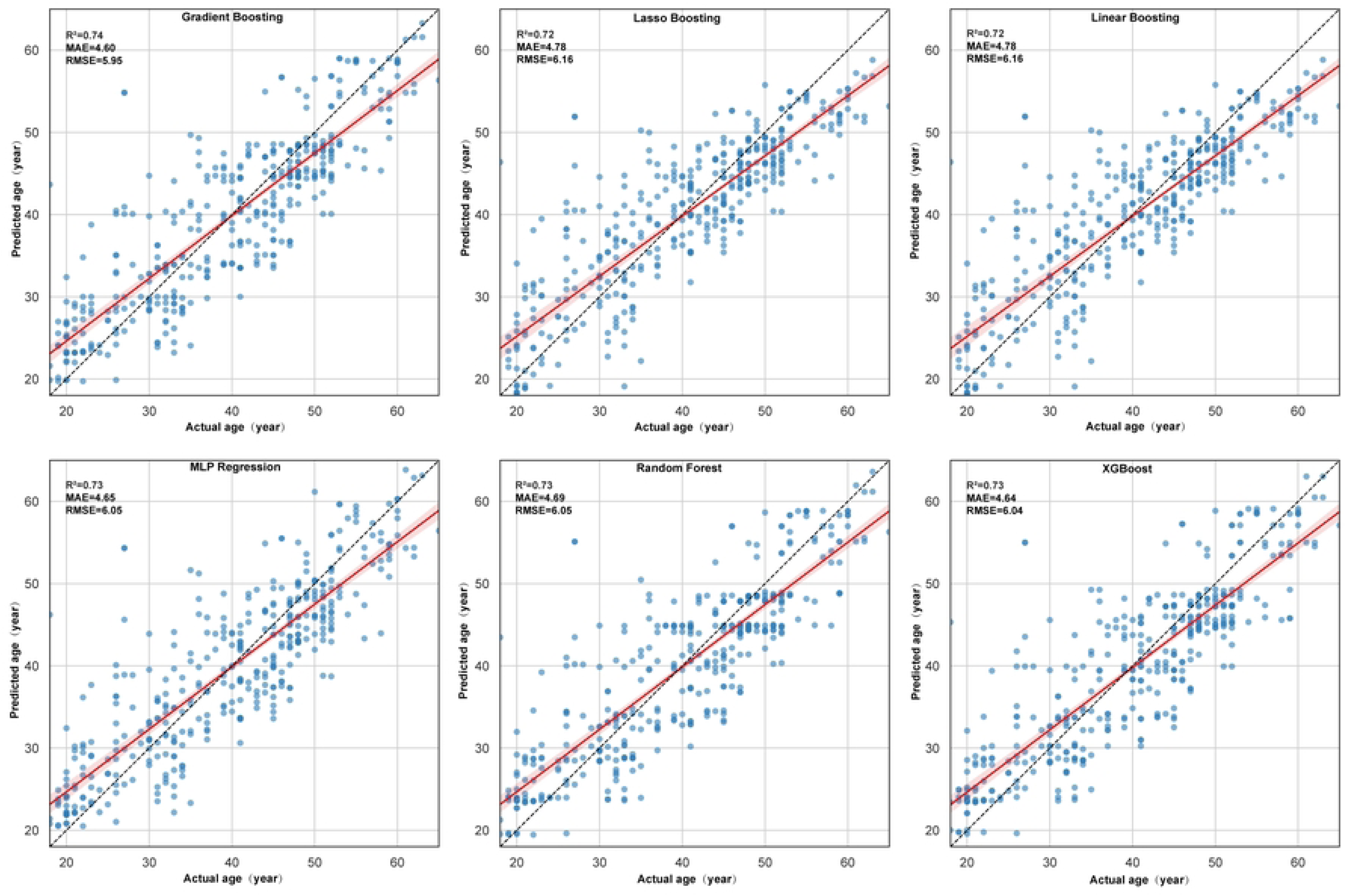
Actual vs. predicted age scatter plots for the six machine learning models in the female sample (position-agnostic but sex-specific models).

**Fig. 7.**
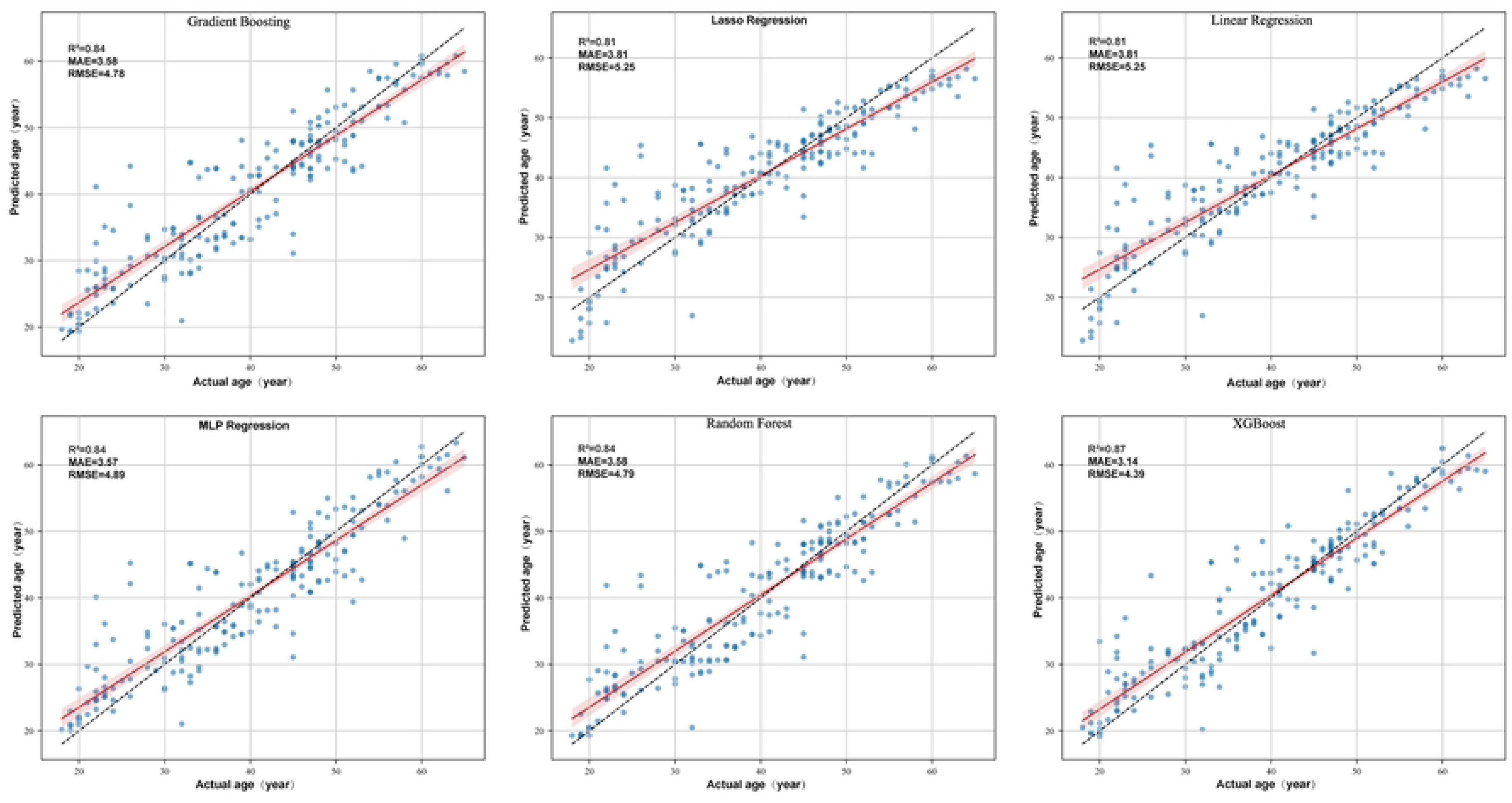
Actual vs. predicted age scatter plots for the six machine learning models in the female sample (position-specific and sex-specific models).

XGBoost consistently achieved the highest performance among all algorithms evaluated within each sex group and the overall sample (Table 6).

## 4. Discussion

Accurate age estimation represents a fundamental task in forensic individual identification, critically influencing the reliability of forensic investigations and judicial outcomes. Traditional skeletal age estimation methods predominantly rely on radiographic assessments of developmental markers in structures such as the clavicle, shoulder, and elbow joints. However, dental age estimation (DAE), leveraging the unique properties of teeth, has garnered considerable research attention due to its superior taphonomic stability and established accuracy. The first molar, as the earliest erupted tooth in the permanent dentition and exhibiting the longest functional duration, demonstrates pronounced age-related morphological alterations, rendering it particularly valuable for age estimation. Previous research has confirmed negligible morphological asymmetry between homonymous contralateral teeth [23]. Therefore, this study employed cone beam computed tomography (CBCT) to perform three-dimensional quantification of pulp chamber volume (PCV) in 1,857 right first molars. This approach aimed to establish a high-precision age estimation model and address a significant research gap within the Han Chinese population.

Traditional two-dimensional radiographic parameters, such as the pulp to tooth area ratio, are inherently limited by projection distortion and anatomical superimposition. These limitations impede their capacity to accurately characterize the complex three-dimensional morphological alterations occurring within the pulp chamber. The present study addressed these limitations by employing high-resolution CBCT ( voxel size ≤200 μm). This technology facilitates the digital reconstruction of the intricate pulp chamber morphology, enabling direct volumetric quantification of PCV. Statistical analysis revealed robust negative correlations between PCV and chronological age (ranging from r = -0.88 to -0.81). The strongest correlation was identified for the maxillary right first molar in females (r = -0.88). This finding aligns with reported sex differences in dental age estimation studies [21, 24], further validating the utility of CBCT-based three-dimensional quantification for capturing the dynamics of secondary dentin deposition.

This study identified statistically significant differences in PCV between maxillary and mandibular first molars [25], with mean PCV being significantly greater in maxillary molars compared to mandibular molars. This finding is consistent with results reported by Ge et al. [18, 26]. This disparity between jaw positions may arise from more regular and distinct incremental growth layers in maxillary teeth compared to their mandibular counterparts [27]. Additionally, mandibular molars typically exhibit more pronounced occlusal wear than their maxillary counterparts. Furthermore, statistically significant sex-based differences in PCV were also observed. This sex-based variation can be explained by two primary factors. First, earlier dental development in females compared to males leads to inherent morphological differences in tooth structure [28]. Second, the differential influence of sex chromosomes on odontogenesis alters mitotic activity within the dental epithelium and papilla [29, 30]. These cellular alterations subsequently result in varying rates of dentin apposition and enamel thinning within the coronal region between sexes. The sex differences observed in the first permanent molars in this study are consistent with the findings of Khamis et al. [31], who reported similar sex-based dental variations across diverse populations, including those from Malaysia, China, Turkey, and Australia. Therefore, in forensic practice, neglecting the variations associated with sex and jaw position may compromise the accuracy of age estimation. Consequently, when confronted with unidentified remains, prior determination of sex and jaw position, followed by application of position- and sex-specific age estimation models, would enhance prediction accuracy and mitigate potential error propagation.

Previous research has consistently demonstrated that the relationship between dental morphological indicators and chronological age frequently exhibits non-linearity [31–33]. For instance, Delphine et al. [31] applied quadratic regression, while Ge et al. [32] and Zhang et al. [33] used logarithmic regression; these studies reported MAE ranging from 6.25 to 9.22 years. Therefore, to identify the optimal regression-based age estimation model for the Sichuan Han Chinese population, this study systematically evaluated eleven curvilinear regression models. Among these models, the cubic regression model demonstrated the highest coefficient of determination (R²) within the training set (R² range: 0.77 - 0.79; Table 4). Specifically, the cubic model utilizing the maxillary right first molar (tooth 16) in females yielded the lowest MAE of 4.95 years (Table 5). However, traditional polynomial regression models exhibit inherent limitations. Their fixed polynomial structure often struggles to adequately accommodate the complex biological variation inherent across different jaw positions and sexes. This limitation is exemplified by the relatively high MAE (6.10 years) observed for the cubic model applied to mandibular molars in males (Table 5). This finding underscores that a single, fixed-form mathematical model may be insufficient for comprehensively capturing the intricate atrophy dynamics within complex biological systems like the dentino-pulpal complex, necessitating more flexible computational approaches.

**Table 5.**
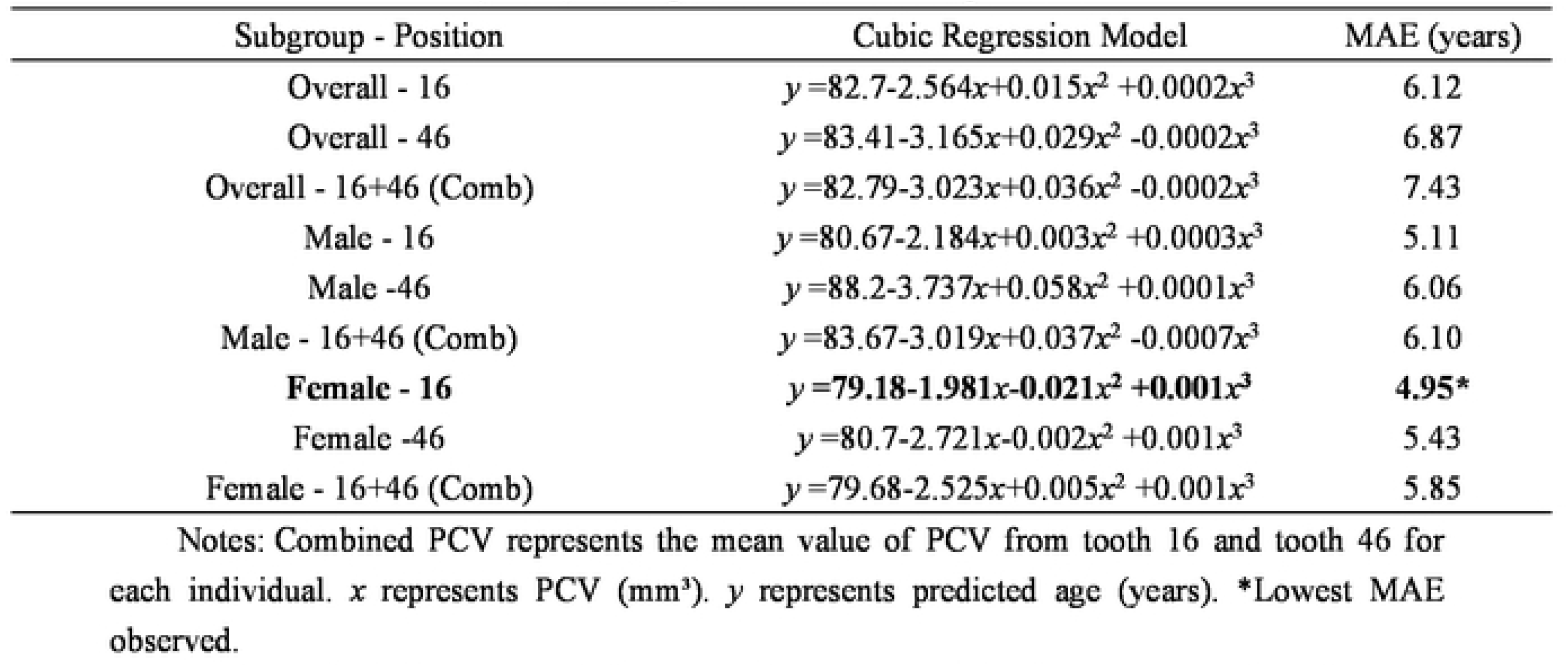
Cubic regression model equations and performance metrics (MAE; years) for age estimation using PCV.

| Subgroup - Position | Cubic Regression Model | MAE (years) |
| --- | --- | --- |
| Overall - 16 | $y = 82.7 - 2.564x + 0.015x^2 + 0.0002x^3$ | 6.12 |
| Overall - 46 | $y = 83.41 - 3.165x + 0.029x^2 - 0.0002x^3$ | 6.87 |
| Overall - 16+46 (Comb) | $y = 82.79 - 3.023x + 0.036x^2 - 0.0002x^3$ | 7.43 |
| Male - 16 | $y = 80.67 - 2.184x + 0.003x^2 + 0.0003x^3$ | 5.11 |
| Male - 46 | $y = 88.2 - 3.737x + 0.058x^2 + 0.0001x^3$ | 6.06 |
| Male - 16+46 (Comb) | $y = 83.67 - 3.019x + 0.037x^2 - 0.0007x^3$ | 6.10 |
| **Female - 16** | $y = 79.18 - 1.981x - 0.021x^2 + 0.001x^3$ | **4.95\*** |
| Female - 46 | $y = 80.7 - 2.721x - 0.002x^2 + 0.001x^3$ | 5.43 |
| Female - 16+46 (Comb) | $y = 79.68 - 2.525x + 0.005x^2 + 0.001x^3$ | 5.85 |

Notes: Combined PCV represents the mean value of PCV from tooth 16 and tooth 46 for each individual. $x$ represents PCV (mm<sup>3</sup>). $y$ represents predicted age (years). \*Lowest MAE observed.

**Table 6.** Performance of best-performing machine learning models for age estimation.

| Sample Group | Model Type | Best Algorithm | R<sup>2</sup> | MAE (years) |
| --- | --- | --- | --- | --- |
| Overall | Position-agnostic | Gradient Boosting | 0.74 | 4.70 |
| Overall | Position-specific | XGBoost | 0.84 | 3.46 |
| Male | Position-agnostic | Gradient Boosting | 0.70 | 4.81 |
| Male | Position-specific | XGBoost | 0.83 | 3.27 |
| Female | Position-agnostic | Gradient Boosting | 0.74 | 4.60 |
| Female | Position-specific | XGBoost | 0.87 | 3.14 |

To overcome the limitations of traditional regression models in capturing the non-linear relationship between PCV and age, this study developed ensemble models incorporating sex and tooth position using six distinct machine learning algorithms. The results demonstrate the following key findings: Significantly Reduced Error in Tooth Position-Specific Models: The tooth position-specific machine learning model achieved the MAE of 3.45 years, representing a significant 26.6% reduction compared to the non-position-specific model (MAE = 4.70 years). This finding further highlights the critical importance of differentiating tooth positions for accurate age estimation.

Superiority of Sex-Specific Models: Sex-specific models outperformed sex-unspecified models. Notably, the female-specific model achieved a lower mean absolute error (MAE = 3.14 years) compared to the male-specific model (MAE = 3.27 years), indicating higher precision for females. These findings may be attributable to the greater temporal stability of dentin mineralization regulated by the X chromosome and the inhibitory effect of estrogen on odontoclast activity.

Overall Superiority of Machine Learning over Traditional Regression Models: The results further confirm the superiority of the developed machine learning models over traditional regression models. This advantage likely stems from two key capabilities inherent to the XGBoost algorithm and its gradient boosting framework: Feature Importance Quantification: Accurately capturing the relative importance of features and their interaction effects, particularly between tooth position and sex. Regularization Techniques: Effectively mitigating overfitting and enabling robust modeling of complex non-linear relationships, such as the decline during the calcification plateau phase observed in older age groups. Consequently, the optimal machine learning model achieved a substantially lower overall MAE of 3.14 years, representing a 36.5% reduction compared to the best-performing traditional cubic regression model.

Furthermore, the performance of our model (MAE = 3.14 years) represents an improvement over the polynomial kernel Support Vector Regression model (reported MAE = 4.86 years) for maxillary lateral incisors developed by Merdietio Boedi et al. [19]. Therefore, the machine learning age estimation models developed in this study, utilizing PCV data from first molars, not only circumvent the limitations of stepwise regression models and facilitate deeper data exploration, but also effectively mitigate the issue of excessive prediction errors associated with low-resolution conventional radiographs. This methodology significantly enhances the accuracy of dental age estimation and offers a novel analytical framework and valuable perspective for future research.

However, this study acknowledges the following three primary limitations: 1. Limited Demographic Scope: The sample cohort was exclusively derived from individuals of Han Chinese ethnicity residing in the Sichuan region, China. To enhance model generalizability, future studies should be expanded to include validation across diverse geographic regions and ethnic groups. 2. Dependence on Semi-automatic Segmentation: The reliance on semi-automatic pulp chamber segmentation currently impedes high-throughput analysis. Future efforts should focus on integrating deep learning algorithms (e.g., Mask R-CNN) to achieve fully automated segmentation. 3. Reliance on a Single Anatomical Feature: The current model relies solely on pulp chamber volume (PCV) as a single predictor. Future research should incorporate multimodal parameters, such as dental root morphological indices and occlusal wear grading, to construct ensemble models. Such integration is essential for developing more accurate and broadly applicable methodologies.

## 5. Conclusion

This study, based on the quantitative analysis of PCV in 1857 first molars from Han Chinese adults using CBCT, confirmed a significant negative correlation between PCV and chronological age (ranging from r = -0.88 to r = -0.81). The strongest correlation was observed in the maxillary first molars of female individuals. The development of sex- and tooth position-specific XGBoost ensemble models reduced the MAE for age estimation to 3.14 years (R² = 0.87), representing a 36.5% improvement in accuracy compared to the best-performing traditional cubic regression model (MAE = 4.95 years). These findings demonstrate that PCV in first molars serves as a valuable indicator for forensic age estimation within the studied Han Chinese population. Future research should focus on enhancing model generalizability and accuracy through: (1) cross-population validation, (2) deep learning-based automated segmentation (e.g., Mask R-CNN), and (3) the integration of additional multimodal dental features.

## Data Availability

Data cannot be shared publicly because of Patient privacy. Data are available from the North Sichuan Medical College Ethics Committee (contact via) for researchers who meet the criteria for access to confidential data

